# Large Language Model-derived Symptom Clusters and Patient Outcomes in Colorectal Cancer from MIMIC-IV Clinical Notes

**DOI:** 10.64898/2026.09.15.26363148

**Authors:** Youran Lee, Ivo Dinov, Xiaosu Hu, Yun Jiang

## Abstract

**Background:** Prior research on symptom clusters (SCs) in colorectal cancer (CRC) has relied primarily on patient-reported outcome surveys, which capture symptom experience at discrete assessment points rather than the continuous documentation generated during routine care, leaving open whether SCs derived from electronic health record (EHR) text carry the same clinical meaning and predictive value. To construct and validate patient-level symptom co-occurrence networks from large language model (LLM)-extracted symptom data in CRC patients, and to test whether resulting SCs predict clinical outcomes.

**Methods:** Using a zero-shot LLM extraction pipeline previously benchmarked against a manually annotated ground truth (Macro F1=0.70 for the best-performing model), we extracted 46 symptoms from 2,728 discharge notes of 1,507 CRC patients in MIMIC-IV. Patient-level symptom co-occurrence networks were constructed independently from Gemini 3.5 Flash and Claude Haiku extractions using phi correlation (≥0.10) and Louvain community detection, with sensitivity analyses across correlation thresholds, random seeds, note-aggregation strategy, and bootstrap resampling. Per-cluster symptom burden scores were tested as predictors of in-hospital mortality, 30-day readmission, and 1-year mortality using logistic regression adjusted for age, sex, and (in sensitivity models) metastatic disease.

**Results:** Both LLMs’ networks converged on three clinically coherent SCs — Systemic, Gastrointestinal, and CRC Disease-Specific — across sensitivity analyses (cross-model Adjusted Rand Index=0.727; 100-seed Louvain ARI=0.985; bootstrap ARI=0.733). The Systemic Symptom Cluster was the most consistent predictor of in-hospital mortality (OR=1.33) and 1-year mortality (OR=1.41), while the CRC Disease-Specific Cluster specifically and independently predicted 30-day readmission (OR=1.20); both associations were robust to adjustment for metastatic disease.

**Conclusion:** LLM-extracted symptom data recover clinically coherent, reproducible SCs from unstructured discharge notes that carry independent prognostic value for mortality and readmission, supporting the clinical validity of automated, EHR-derived symptom profiling in CRC.

**Highlights:**

- LLMs identified Systemic, CRC Disease-Specific, and Gastrointestinal clusters.
- Three symptom clusters were reproducible across LLMs and sensitivity analyses.
- Systemic symptom burden predicted in-hospital and 1-year mortality.

## 1. Introduction

Colorectal cancer (CRC), the third-leading cause of cancer deaths [1], is associated with persistent physical and psychological symptoms that occur in clusters and are interrelated (i.e., symptom clusters, SCs) [2]. Patients with CRC often experience multiple concurrent symptoms, including fatigue, pain, gastrointestinal symptoms, sleep disturbance, and psychological distress, across the disease trajectory [2, 3]. These SCs have synergistic and cumulative effects that negatively affect patient quality of life, treatment adherence, and healthcare outcomes [4–6]. Understanding how symptoms co-occur is therefore important for identifying clinically meaningful patterns of symptom burden and patients who may be at increased risk for adverse outcomes.

Most knowledge about SCs in CRC has been generated from studies using patient-reported outcome measures [7]. Electronic health records (EHRs), however, provide an additional source of symptom information generated during routine clinical care, much of which is embedded in free-text clinical narratives rather than structured fields [8].

Previous studies have demonstrated the feasibility of extracting symptoms from these narratives using natural language processing (NLP), including constructing symptom co-occurrence networks and identifying SCs among patients with cancer [9]. However, conventional approaches may require substantial preprocessing, manually developed rules, annotated training data, or task-specific model development [10]. Large language models (LLMs) offer a scalable alternative for transforming unstructured clinical narratives into structured symptom data [11, 12]. Although recent studies have demonstrated the accuracy of LLM-based clinical information extraction and its potential for downstream prediction, whether LLM-derived symptom data can support clinically meaningful network-based symptom phenotyping in CRC and whether the resulting SCs are associated with subsequent clinical outcomes remain unclear.

Our prior work demonstrated that zero-shot LLMs can accurately extract CRC symptoms from unstructured clinical notes, establishing the feasibility of generating structured symptom data at scale [13]. The next critical step is to determine whether these extracted symptoms can be translated into clinically meaningful symptom phenotypes and whether these phenotypes are associated with subsequent clinical outcomes. Therefore, this study aimed to (1) construct symptom co-occurrence networks and identify SCs among patients with CRC and (2) evaluate the clinical validity of these SCs by examining their associations with in-hospital mortality, 30-day readmission, and 1-year mortality. By linking automated symptom extraction with network-based symptom phenotyping and downstream outcome evaluation, this study examines the potential of routinely documented clinical narratives to support scalable and clinically meaningful symptom profiling in CRC.

## 2. Methods

### 2.1 Data Source

This study utilized the MIMIC-IV Clinical Notes dataset (version 3.1), a de-identified clinical note database from Beth Israel Deaconess Medical Center available through PhysioNet [14]. Discharge summaries — which synthesize the clinical course and symptom burden of an entire admission in narrative form — were extracted for patients diagnosed with CRC, identified using ICD-9 codes (153.x, 154.x) and ICD-10 codes (C18–C20) from the MIMIC-IV hospital module. A total of 1,507 patients and 2,728 discharge notes were included in the analysis.

### 2.2 Symptom Extraction

Symptom presence was extracted from the Chief Complaint and History of Present Illness (CC/HPI) sections of each discharge note using a zero-shot LLM pipeline (Gemini 3.5 Flash; Claude Haiku used for cross-model robustness checks), applied to a target list of 46 symptoms derived from the Memorial Symptom Assessment Scale [15] and the EORTC QLQ-CR29 [16]. Both models were selected based on benchmarking against a manually annotated, adjudicated 200-note gold standard, reported in a companion study (Gemini 3.5 Flash Macro F1=0.70; Claude Haiku Macro F1=0.63; both substantially outperforming rule-based and NER baselines) [13]. Full extraction methodology, prompts, and ground-truth annotation procedures are reported in that companion study.

### 2.3 Symptom Network Analysis

To examine co-occurrence patterns among symptoms, a patient-level symptom co-occurrence network was constructed, consistent with established network analytic approaches for examining relationships among symptoms [17]. Binary symptom indicators extracted by each LLM (Gemini 3.5 Flash and Claude Haiku) were independently aggregated at the patient level: a symptom was coded as present (1) if documented in any discharge note for a given patient, and absent (0) otherwise. Pairwise phi correlations were calculated between all symptom pairs across the 1,507 patients to quantify co-occurrence strength. Symptoms with prevalence below 5% were excluded to improve network stability; this threshold retained 20 of 46 symptoms for Gemini and 21 of 46 for Claude. Excluded symptoms and their prevalence rates are presented in Supplementary Materials.

In the resulting network, symptoms served as nodes and weighted edges represented phi correlations ≥0.10, with node size scaled to prevalence and edge thickness to correlation strength. Strength centrality (sum of absolute edge weights per node) was used to identify the most influential symptoms [17]. No negative phi correlations exceeded the ϕ≥0.10 threshold in either network (Gemini range: 0.006–0.760; Claude minimum: −0.005), so centrality estimates were unaffected. SCs were identified using Louvain community detection (random_state=42), run independently for each LLM’s predictions to enable direct comparison. Following established principles of sensitivity analysis [18], four pre-specified sensitivity analyses assessed: (1) cluster stability across phi thresholds (0.10, 0.15, 0.20); (2) assignment consistency across 100 random Louvain seeds via adjusted Rand index (ARI); (3) whether restricting to each patient’s first discharge note (vs. any-note aggregation) inflates prevalence estimates; and (4) cluster stability under 200 bootstrap resamples via co-assignment proportions and ARI. Analyses used Python’s networkx and python-louvain libraries.

### 2.4 Predictive Validity Analysis

To evaluate predictive validity, logistic regression models were fitted to examine the associations between per-cluster symptom burden scores and binary clinical outcomes [19]. Per-cluster symptom burden scores, defined as the count of symptoms present within each cluster and derived from the Gemini 3.5 Flash-based network, were evaluated as predictors of three clinical outcomes: in-hospital mortality, 30-day readmission, and 1-year mortality (N=1,507). All three cluster burden scores were entered simultaneously as continuous predictors, with adjustment for age and sex; a post hoc sensitivity analysis additionally adjusted for metastatic disease. Outcomes were defined consistently with Table 1: in-hospital mortality as death during any admission, 30-day readmission as any rehospitalization within 30 days of the first discharge, and 1-year mortality as death within one year of the first admission.

**Table 1.** Demographic Characteristics of the Study Cohort (N = 1,507)

| Characteristic | N = 1,507 |
| --- | --- |
| Age, mean ± SD (median [IQR]), years | 65.5 ± 15.0 (66 [54–77]) |
| Sex, n (%) |  |
| Male | 774 (51.4) |
| Female | 733 (48.6) |
| Race/ethnicity, n (%) |  |
| White | 1,039 (68.9) |
| Black/African American | 185 (12.3) |
| Asian | 94 (6.2) |
| Hispanic/Latino | 43 (2.9) |
| Other/Unknown | 146 (9.7) |
| Insurance, n (%) |  |
| Medicare | 723 (48.0) |
| Private | 522 (34.6) |
| Medicaid | 218 (14.5) |
| Other/No charge | 44 (2.9) |
| Marital status |  |
| Married/Partnered | 736 (48.8) |
| Single | 449 (29.8) |
| Divorced/Separated | 91 (6.0) |
| Widowed | 203 (13.5) |
| Unknown | 28 (1.9) |
| **CRC Characteristics** |  |
| **Tumor site (ICD-coded), n (%)<sup>a</sup>** |  |
| Colon (C18 / ICD-9 153.x) | 1,020 (67.7) |
| Rectosigmoid (C19 / ICD-9 154.5) | 107 (7.1) |
| Rectum (C20 / ICD-9 154.x) | 495 (32.8) |
| Metastatic disease (any admission), n (%) | 775 (51.4) |
| **Treatment (procedure-coded)** |  |
| Surgical resection (any admission), n (%) | 941 (62.4) |
| Chemotherapy (any admission), n (%) | 113 (7.5) |
| Radiation therapy (any admission), n (%) | 71 (4.7) |
| **Comorbidity** |  |
| Charlson Comorbidity Index, mean ± SD (median [IQR])<sup>b</sup> | 6.0 ± 3.0 (6 [3–8]) |
| CCI 1–2, n (%) | 252 (16.7) |
| CCI 3–4, n (%) | 242 (16.1) |
| CCI ≥5, n (%) | 1,013 (67.2) |
| **Hospitalization** |  |
| No. of hospital admissions per patient, mean ± SD (median) | 4.4 ± 4.2 (3) |
| Length of stay per admission, mean ± SD (median), days | 5.4 ± 6.3 (3.7) |
| Surgical admission type (any), n (%) | 940 (62.4) |
| Emergency or urgent admission (any), n (%) | 1,128 (74.9) |
| **Clinical Outcomes** |  |
| In-hospital mortality (any admission), n (%) | 155 (10.3) |
| 30-day readmission (from first discharge), n (%) | 409 (27.1) |
| 1-year mortality (from first admission), n (%) | 233 (15.5) |

*Note.* Age reported as mean ± SD and median [IQR]. IQR = interquartile range; SD = standard deviation; CCI = Charlson Comorbidity Index. <sup>a</sup>Tumor site categories are not mutually exclusive; patients with synchronous tumors may appear in multiple categories. Metastatic disease defined by presence of C77–C80 (ICD-10) or 196–199 (ICD-9) codes in any admission. <sup>b</sup>CCI computed using Quan et al. (2005) mapping applied to both ICD-9 and ICD-10 diagnosis codes; categories are not mutually exclusive across ICD version. All cancer patients have CCI ≥ 2 (primary cancer weight). Treatment status derived from ICD-9/10 procedure codes across all CRC admissions. Clinical outcomes derived from MIMIC-IV admissions and patient records.

## 3. Results

### 3.1 Patient Characteristics

Table 1 summarizes the characteristics of the 1,507 patients with CRC. The mean age was 65.5 years (SD 15.0), 51.4% were male, and 51.4% had metastatic disease. Comorbidity burden was high (mean CCI 6.0, SD 3.0), and 74.9% had an emergency or urgent admission. In-hospital mortality was 10.3% (n=155), 30-day readmission was 27.1% (n=409), and 1-year mortality was 15.5% (n=233).

### 3.2 Symptom Cluster Analysis

Symptom network analyses were conducted separately using symptom predictions from Gemini 3.5 Flash and Claude Haiku to assess the consistency of identified clusters across extraction methods, applying identical parameters: phi correlation threshold ≥0.10, prevalence threshold ≥5%, and Louvain community detection (random_state=42).

Gemini 3.5 Flash-Based Network. After excluding 26 symptoms with prevalence below 5%, 20 symptoms were retained for network construction (phi range: 0.005–0.759). The Louvain algorithm identified three distinct SCs (Figure 1, Figure 2, Table 2).

**Figure 1.**
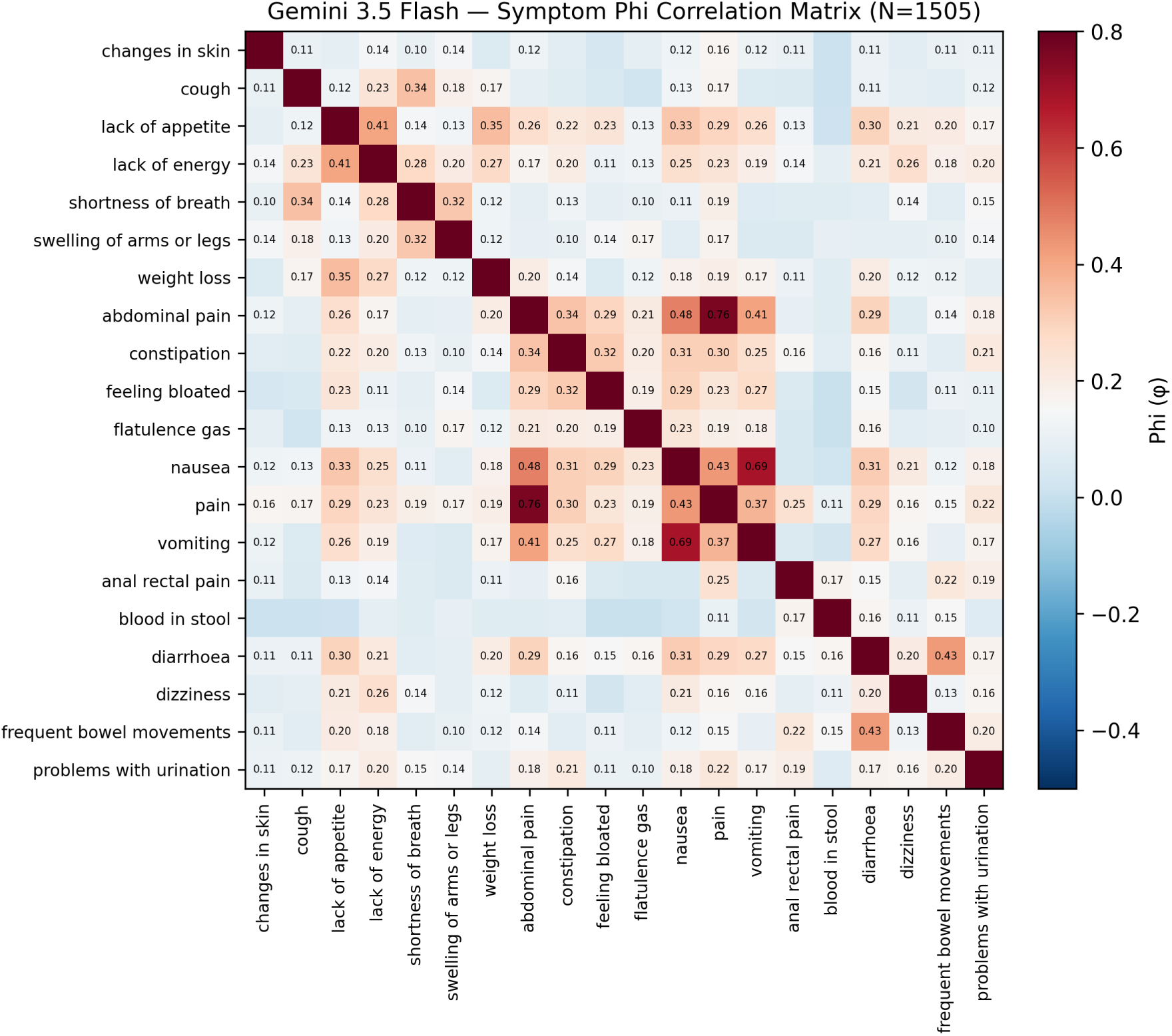
Symptom Phi Correlation Matrix (Gemini 3.5 Flash; N=1,507 CRC Patients) Note: Phi correlation between each pair of 20 symptoms (prevalence ≥5%) across 1,507 patients. Color scale: blue = negative correlation, white ≈ zero, red = strong positive correlation. Rows and columns are sorted by Louvain cluster membership.

**Figure 2a.**
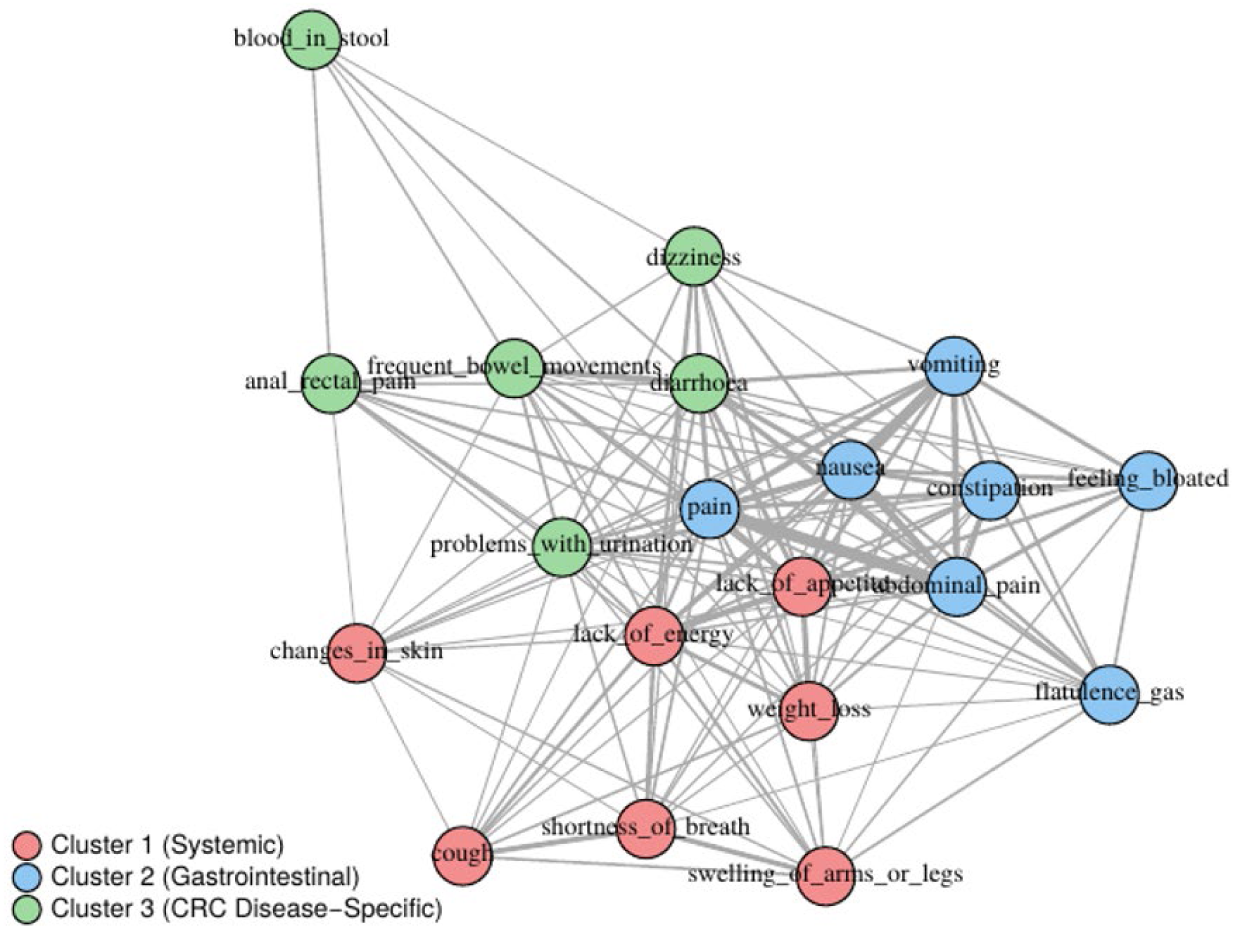
Symptom Co-occurrence Network *— Gemini 3.5 Flash*.

**Figure 2b.**
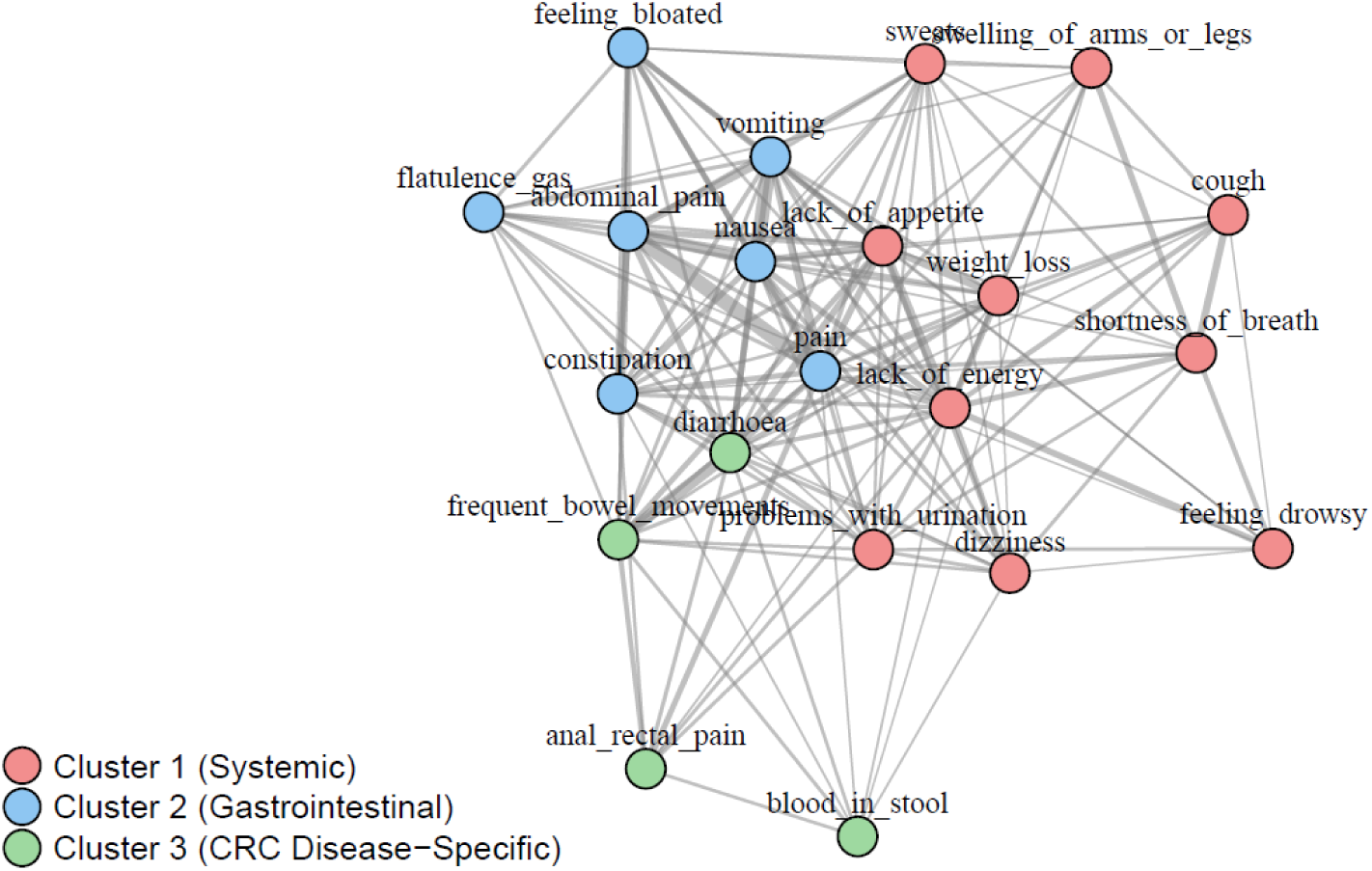
Symptom Co-occurrence Network — Claude Haiku. Note: Nodes represent symptoms; node size is proportional to prevalence. Edge thickness reflects phi correlation strength (phi ≥ 0.10 shown). Colors denote Louvain clusters (see Table 2 for A/B/C codes).

**Table 2.** Symptom Clusters Identified by Louvain Community Detection (Gemini 3.5 Flash; N=1,507 CRC Patients)

| Code | Symptom | Cluster | Prevalence (%) | Strength Centrality |
| --- | --- | --- | --- | --- |
| A1 | Lack of Appetite | 1 | 23.0 | 3.88 |
| A2 | Lack of Energy | 1 | 25.4 | 3.81 |
| A3 | Weight Loss | 1 | 18.5 | 2.58 |
| A4 | Shortness of Breath | 1 | 16.2 | 2.14 |
| A5 | Swelling of Arms or Legs | 1 | 8.2 | 1.92 |
| A6 | Cough | 1 | 9.2 | 1.68 |
| A7 | Changes in Skin | 1 | 6.8 | 1.45 |
| **B1** | Diarrhea | 2 | 23.8 | 3.7 |
| **B2** | Problems with Urination | 2 | 11.9 | 2.8 |
| **B3** | Frequent Bowel Movements | 2 | 8.4 | 2.35 |
| **B4** | Dizziness | 2 | 11.3 | 1.96 |
| **B5** | Anal Rectal Pain | 2 | 6.0 | 1.63 |
| **B6** | Blood in Stool | 2 | 29.1 | 0.7 |
| **C1** | Pain | 3 | 50.9 | 4.86 |
| **C2** | Nausea | 3 | 25.1 | 4.38 |
| **C3** | Abdominal Pain | 3 | 37.4 | 3.84 |
| **C4** | Vomiting | 3 | 19.8 | 3.51 |
| **C5** | Constipation | 3 | 17.9 | 3.15 |
| **C6** | Feeling Bloated | 3 | 10.9 | 2.44 |
| **C7** | Flatulence Gas | 3 | 5.6 | 2.11 |

*Note. Strength centrality = sum of absolute phi correlations for each node. Prevalence = proportion of patients with symptom present in any discharge note. Symptoms with prevalence <5% were excluded prior to network construction. $\Phi \geq 0.10$ threshold applied for edge inclusion. Clusters 1–3 correspond to Systemic, CRC Disease-Specific, and Gastrointestinal Symptom Clusters, respectively.*

*Cluster 1:* Systemic Symptom Cluster (n=7 symptoms) comprised lack of appetite, lack of energy, weight loss, shortness of breath, swelling of arms or legs, cough, and changes in skin, with lack of appetite showing the highest strength centrality (3.88).

*Cluster 2:* CRC Disease-Specific Symptom Cluster (n=6 symptoms) included diarrhea, problems with urination, frequent bowel movements, dizziness, anal rectal pain, and blood in stool, with diarrhea showing the highest strength centrality (3.70), consistent with its central role in CRC symptomatology.

*Cluster 3:* Gastrointestinal Symptom Cluster (n=7 symptoms) comprised pain, nausea, abdominal pain, vomiting, constipation, feeling bloated, and flatulence/gas. Pain was both the most prevalent symptom overall (50.9%) and, along with nausea, showed the highest strength centrality (4.86 and 4.38, respectively).

Claude Haiku-Based Network. An identical analysis on Claude Haiku symptom predictions yielded 21 symptoms meeting the prevalence threshold and again identified three distinct SCs with a broadly similar structure (Figure 2 and Table 2).

*Cluster 1:* Systemic Symptom Cluster (n=10 symptoms) additionally comprised problems with urination, sweats, dizziness, and feeling drowsy relative to the Gemini network — feeling drowsy (10.2%) and sweats (7.1%) appeared here owing to their higher documented prevalence in Claude-extracted data.

*Cluster 2:* CRC Disease-Specific Symptom Cluster (n=4 symptoms) included diarrhea, frequent bowel movements, anal rectal pain, and blood in stool — the most compact cluster of the three, excluding dizziness and problems with urination unlike the Gemini network.

*Cluster 3:* Gastrointestinal Symptom Cluster (n=7 symptoms) was identical in membership to the Gemini Gastrointestinal Symptom Cluster, the most stable grouping across both models.

Cross-Model Comparison. Both Gemini and Claude network analyses converged on three clinically coherent SCs — Systemic, CRC Disease-Specific, and Gastrointestinal — supporting the robustness of this symptom organization across extraction methods. The Gastrointestinal Symptom Cluster was identical across both models, and weight loss was assigned to the Systemic cluster by both. Structural differences were confined to the Systemic and CRC Disease-Specific clusters, reflecting redistribution of a few boundary symptoms (sweats, feeling drowsy, dizziness, problems with urination) rather than a different underlying organization — suggesting that symptom burden in CRC patients is consistently organized into distinct physiological axes across LLM extraction methods.

### 3.3 Sensitivity Analyses

Phi correlation threshold sensitivity. A three-cluster solution was obtained at both ϕ≥0.10 (136 edges) and ϕ≥0.15 (88 edges), but cluster membership shifted substantially between thresholds (ARI=0.438; cluster sizes 7/7/6 vs. 11/5/4), indicating the three-community structure was preserved in number but not composition as weaker edges were pruned. At ϕ≥0.20 (51 edges), the network fragmented into five clusters as weakly-connected symptoms (changes in skin, blood in stool) became isolated singletons. These results support ϕ≥0.10 as a threshold that preserves clinically meaningful structure while avoiding over-connectivity.

Louvain cluster stability. Across 100 random seeds at ϕ≥0.10, the Louvain algorithm consistently identified three clusters in all iterations, with high assignment consistency relative to the reference partition (seed=42; mean ARI=0.985, range 0.850–1.000); deviations reflected re-labeling of boundary symptoms across cluster borders rather than fundamental structural change.

Bootstrap cluster stability (patient resampling). Across 200 bootstrap resamples of the 1,507 patients, Louvain community detection recovered the three-cluster solution in 192 iterations (96.0%); mean ARI relative to the reference partition was 0.733 (SD=0.159, range: 0.349–1.000, median: 0.724), indicating moderate-to-high cluster assignment consistency across samples.

First-note vs. any-note aggregation. Restricting symptom aggregation to each patient’s first discharge note yielded lower mean symptom burden (2.73 symptoms/patient) than any-note aggregation (4.15 symptoms/patient); several symptoms fell below the 5% prevalence threshold under first-note restriction, including frequent bowel movements (8.4%→4.8%), swelling of arms or legs (8.2%→4.3%), changes in skin (6.8%→3.5%), anal rectal pain (6.0%→3.8%), and flatulence/gas (5.6%→3.3%). This suggests any-note aggregation captures cumulative symptom burden across the care episode but may include symptoms that resolved before the final discharge note; future studies should consider time-stamped sensitivity analyses to account for symptom temporality.

### 3.4 Predictive Validity of Symptom Clusters

In-hospital mortality (10.3%, n=155). Higher Systemic Symptom Cluster burden was significantly associated with in-hospital mortality (OR=1.33 per symptom, 95% CI: 1.18– 1.49, p<0.001). Gastrointestinal Symptom Cluster burden was also associated (OR=1.13, 95% CI: 1.02–1.25, p=0.015; not retained after stage adjustment, see below), whereas CRC Disease-Specific Cluster burden was not (OR=1.01, 95% CI: 0.87–1.17, p=0.873), after simultaneous adjustment for all three cluster burdens, age, and sex.

30-day readmission (27.1%, n=409). CRC Disease-Specific Cluster burden was the only significant predictor of 30-day readmission (OR=1.20, 95% CI: 1.08–1.34, p<0.001). Neither Systemic Cluster burden (OR=1.09, 95% CI: 0.99–1.19, p=0.067) nor Gastrointestinal Symptom Cluster burden (OR=1.02, 95% CI: 0.96–1.10, p=0.496) was independently associated with readmission after adjustment for age and sex.

1-year mortality (15.5%, n=233). Systemic Symptom Cluster burden was the strongest predictor of 1-year mortality (OR=1.41, 95% CI: 1.27–1.56, p<0.001), followed by Gastrointestinal Cluster burden (OR=1.11, 95% CI: 1.02–1.21, p=0.018). CRC Disease-Specific Cluster burden was inversely associated with 1-year mortality (OR=0.85, 95% CI: 0.74–0.98, p=0.023).

Post hoc sensitivity analysis (metastatic disease adjustment). Because the inverse association between CRC Disease-Specific Cluster burden and 1-year mortality could reflect confounding by disease stage, all models were refitted with additional adjustment for metastatic disease (n=775, 51.4%), itself a strong predictor of 1-year mortality (OR=4.57, 95% CI: 3.17–6.58, p<0.001). The inverse CRC Disease-Specific association persisted and strengthened (OR=0.83, 95% CI: 0.71–0.95, p=0.009), as did the Systemic association (OR=1.32, 95% CI: 1.19–1.48, p<0.001). Metastatic disease was more, not less, prevalent among patients with higher CRC Disease-Specific burden (43.5% at burden 0 rising to 78.8% at burden 3), and the inverse association was confined to patients with metastatic disease (OR=0.81, 95% CI: 0.69–0.95, p=0.009 vs. OR=0.93, 95% CI: 0.65–1.32, p=0.679 without) — arguing against a less-advanced-disease explanation. Gastrointestinal Cluster burden lost significance for both 1-year mortality (OR=1.01, 95% CI: 0.92–1.11, p=0.817) and in-hospital mortality (OR=1.08, 95% CI: 0.97–1.19, p=0.148) after adjustment, indicating its signal is partly explained by disease stage. Readmission estimates were unchanged (CRC Disease-Specific OR=1.20, 95% CI: 1.08–1.34, p<0.001). Overall, Systemic and CRC Disease-Specific cluster associations remained robust after adjustment for metastatic disease, whereas Gastrointestinal cluster associations with mortality did not.

## 4. Discussion

To our knowledge, this is the first study to identify symptom clusters in patients with colorectal cancer using LLM-extracted symptom data from EHR clinical narratives. In this study of 1,507 CRC patients with substantial comorbidity burden and high rates of adverse outcomes (10.3% in-hospital mortality, 27.1% 30-day readmission, 15.5% 1-year mortality), network analysis of LLM-extracted symptoms — using an extraction method previously benchmarked against manual annotation (Macro F1=0.70) [13] — consistently identified three clinically coherent SCs (Systemic, CRC Disease-Specific, and Gastrointestinal) across both LLMs and multiple sensitivity analyses, with the Gastrointestinal cluster showing the greatest stability between models. These clusters also showed predictive validity: after adjustment for age and sex, the Systemic cluster was the most consistent prognostic indicator across both mortality outcomes, while the CRC Disease-Specific cluster specifically predicted 30-day readmission; both were robust to further adjustment for metastatic disease, whereas the Gastrointestinal cluster’s mortality association was not, indicating it largely reflects disease stage rather than an independent contribution.

These findings both converge with and diverge from the broader CRC symptom cluster literature. A recent systematic review demonstrated substantial heterogeneity in cluster composition across assessment instruments and analytic methods, while also identifying several recurring patterns, including fatigue–psychological, nausea–vomiting–appetite loss, and abdominal pain–bloating clusters [20]. Recent network-based studies similarly identified CRC-specific, gastrointestinal, and energy-related symptom clusters, with bowel habit changes emerging as a central CRC-specific symptom [21]. These patterns broadly parallel the Systemic, Gastrointestinal, and CRC Disease-Specific SCs identified in our study, despite substantial differences in symptom ascertainment and clinical setting. Notably, prior work has also demonstrated that symptom clusters can be derived directly from CRC clinical narratives using conventional NLP and graph-based methods, providing complementary evidence that clinically meaningful symptom relationships are represented in routinely documented EHR text [22].

Important differences also emerged in cluster composition. Psychological symptoms, which commonly cluster with fatigue and systemic symptoms in patient-reported CRC studies [20, 23], were not prominent in our LLM-derived networks. This may reflect both documentation and extraction differences: clinician-authored discharge notes may prioritize acute physical complaints, and LLM-derived psychological symptom estimates have shown only modest correspondence with contemporaneous patient-reported measures [24]. However, a prior CRC EHR study using MetaMap identified an anxiety– insomnia–weakness–depression cluster [22], suggesting that psychological symptom representation may depend on documentation source and extraction strategy. Direct comparisons of LLM-extracted symptoms with contemporaneous PROs are needed to distinguish documentation bias from extraction error.

The inverse association between CRC Disease-Specific Cluster burden and 1-year mortality warrants careful interpretation. One potential explanation—confounding by disease stage, whereby patients with localized bowel symptoms may have less advanced disease—was not supported by our findings: metastatic disease was more common among patients with higher CRC Disease-Specific burden, and the inverse association strengthened after adjustment for metastatic disease and remained evident within the metastatic subgroup. One hypothesis is that documented organ-specific bowel symptoms may characterize patients whose disease burden remains expressed as discrete, clinically actionable complaints, consistent with this cluster also being the only significant predictor of 30-day readmission. In contrast, constitutional symptoms may reflect more generalized systemic decline. Documentation bias may also contribute to this pattern.

Recent large-scale EHR evidence demonstrates substantial discrepancies between patient-reported symptoms and clinician documentation, with most symptoms reported more frequently by patients and generally low-to-moderate agreement between the two sources [25]. Thus, the absence of documentation of specific bowel symptoms may not necessarily indicate symptom absence or resolution. Because the present analysis was cross-sectional and based on symptom presence rather than timing or trajectory, these interpretations remain hypotheses requiring prospective validation.

This study has several limitations. First, symptom aggregation across all discharge notes does not account for temporality. Restricting the analysis to each patient’s first discharge note resulted in lower symptom burden, suggesting that multi-note aggregation captures cumulative symptom burden but may also include symptoms that had resolved over the course of care. Second, the phi correlation (≥0.10) and prevalence (≥5%) thresholds were prespecified rather than empirically optimized, although sensitivity analyses demonstrated the robustness of the cluster structure across alternative phi thresholds.

Third, clusters were derived using a variable-centered approach that identifies symptoms that co-occur across the cohort and were linked to outcomes using unweighted cluster burden scores. This differs conceptually from person-centered approaches such as latent class analysis, which identify subgroups of patients with similar symptom profiles. Future studies could directly compare network-based symptom clusters with person-centered phenotypes derived from the same EHR symptom data. Finally, this study was conducted using data from a single academic medical center, and external validation across health systems and clinical settings is needed to establish generalizability.

These findings support LLM-based symptom extraction as a scalable approach to symptom surveillance from routinely collected clinical narratives [11]. The identified SCs—particularly the CRC Disease-Specific cluster characterized by diarrhea, blood in stool, and anal/rectal pain—may help inform targeted symptom assessment and patient-reported outcome monitoring. An important next step is to move beyond cross-sectional co-occurrence toward longitudinal symptom dynamics. Recent CRC studies have identified core and sentinel symptoms within symptom networks [21] and shown that symptom burden and cluster structure can evolve across chemotherapy cycles [26]. Applying longitudinal approaches to sequential EHR notes could identify temporal antecedent or “sentinel” symptoms, extending LLM-derived symptom profiling toward prospective early-warning indicators for symptom management.

## Ethics and Data Availability

Ethics statement: This study used the MIMIC-IV database under a valid data use agreement (DUA) with PhysioNet. The study protocol was submitted to the University of Michigan Medicine Institutional Review Board, which determined that the study was not regulated. This determination reflects the study’s exclusive use of the publicly available, fully de-identified MIMIC-IV dataset, with no direct interaction with or identifiable information about human subjects.

## Data availability

The MIMIC-IV dataset is publicly available at https://physionet.org/content/mimiciv/ to credentialed researchers who complete the required training and data use agreement.

## Code availability

All extraction pipelines, annotation guidelines, and analysis scripts are publicly available at https://github.com/youranrl-dot/symptom-cluster_networking_analysis.

## Author contributions

Y.L.: Conceptualization, Methodology, Software, Formal analysis, Writing — original draft. I.D.: Methodology, Formal analysis, Writing — review and editing. X.H.: Methodology, Software, Formal analysis: Supervision, Writing — review and editing.

